# COVID-19 vaccine perceptions: An observational study on Reddit

**DOI:** 10.1101/2021.04.09.21255229

**Authors:** Navin Kumar, Isabel Corpus, Meher Hans, Nikhil Harle, Nan Yang, Curtis McDonald, Shinpei Nakamura Sakai, Kamila Janmohamed, Weiming Tang, Jason L. Schwartz, S. Mo Jones-Jang, Koustuv Saha, Shahan Ali Memon, Chris T. Bauch, Munmun De Choudhury, Orestis Papakyriakopoulos, Joseph D. Tucker, Abhay Goyal, Aman Tyagi, Kaveh Khoshnood, Saad Omer

## Abstract

**Objectives:** As COVID-19 vaccinations accelerate in many countries, narratives skeptical of vaccination have also spread through social media. Open online forums like Reddit provide an opportunity to quantitatively examine COVID-19 vaccine perceptions over time. We examine COVID-19 misinformation on Reddit following vaccine scientific announcements.

**Methods:** We collected all posts on Reddit from January 1 2020 - December 14 2020 (n=266,840) that contained both COVID-19 and vaccine-related keywords. We used topic modeling to understand changes in word prevalence within topics after the release of vaccine trial data. Social network analysis was also conducted to determine the relationship between Reddit communities (subreddits) that shared COVID-19 vaccine posts, and the movement of posts between subreddits.

**Results:** There was an association between a Pfizer press release reporting 90% efficacy and increased discussion on vaccine misinformation. We observed an association between Johnson and Johnson temporarily halting its vaccine trials and reduced misinformation. We found that information skeptical of vaccination was first posted in a subreddit (r/Coronavirus) which favored accurate information and then reposted in subreddits associated with antivaccine beliefs and conspiracy theories (e.g. conspiracy, LockdownSkepticism).

**Conclusions:** Our findings can inform the development of interventions where individuals determine the accuracy of vaccine information, and communications campaigns to improve COVID-19 vaccine perceptions. Such efforts can increase individual- and population-level awareness of accurate and scientifically sound information regarding vaccines and thereby improve attitudes about vaccines. Further research is needed to understand how social media can contribute to COVID-19 vaccination services.

**Funding:** Study was funded by the Yale Institute for Global Health and the Whitney and Betty MacMillan Center for International and Area Studies at Yale University. The funding bodies had no role in the design, analysis or interpretation of the data in the study.

## Introduction

Coronavirus Disease 2019 (COVID-19) vaccine development and widespread scale-up are major steps in combating the pandemic [1]. Several vaccine candidates appear to provide protection not just against disease but also against infection [2], meaning the vaccines could be instrumental in significantly reducing infections among populations. However, for vaccines to be successful, they not only need to be deemed safe and effective by scientists, but also widely accepted by the public [3]. Effective health communication is key to vaccine acceptance, but is a complex task given widespread vaccine hesitancy, rapidly changing vaccine information [4, 5], and vaccine misinformation [6, 7]. Misinformation is defined as information that has the features of being false, determined based on expert evidence, but shared with no intention of harm [8]. For example, some falsely believe that participants in vaccine trials have died after taking the vaccine, or that vaccination is a means to covertly implant microchips [9, 10]. Such information may worsen existing fear around a vaccine and limit public uptake of a COVID-19 vaccine [3]. There are a diverse range of individuals who are skeptics of vaccination including those who are antivaccine or antivaxxers (individuals who are opposed to vaccination or laws that require vaccination) and those who are vaccine hesitant (those who delay in acceptance or refusal of vaccination) [11]. The diverse groups who are skeptical of vaccines may react to information in different ways [12]. With low willingness to vaccinate globally [13], and substantial COVID-19 misinformation [14], achieving sufficient vaccination coverage to reach population-level benefits will be challenging.

Reduced vaccine uptake may impinge on population-level impact [15], and COVID-19 control at the population level [16]. For example, reduced vaccine uptake may increase the mortality cost of COVID-19 [16] and create clusters of non-vaccinators that disproportionately increase pandemic spread [17]. In addition, willingness to accept a COVID-19 vaccine seems to be fluctuating in the US [18, 19], with decreases in vaccine acceptance leading up to the 2020 Presidential Election in the United States (US), perhaps due to significant politicization of the vaccine [19]. Thus, vaccine acceptance is not constant or uniform, and likely affected by several factors, such as being responsive to information and perceptions regarding the vaccine, and the state of the pandemic and economy.

Several studies have detailed the relationship between exposure to COVID-19 misinformation and vaccine acceptance [3], as well as COVID-19 vaccine perceptions assessed via Twitter [6, 20] and online surveys [21, 22, 23, 24, 25]. However, limited research has explored how online vaccine perceptions are associated with major events in the vaccine development and implementation timeline (e.g. major pharmaceutical firms halting vaccine trials or publishing results on vaccine effectiveness) and how online vaccine discussions move across arenas that have different baseline vaccine perceptions.

To our knowledge, prior studies have generally not focused on Reddit, a social news aggregation and discussion website. Registered Reddit members submit posts (text, images, videos) to the site, which are then voted up or down by other members. Posts are organized by subject into user-created boards called communities or subreddits, which cover a large range of topics. Reddit may be a useful setting for examining vaccine perceptions because similar topics have been discussed before [26], including topics related to COVID-19 vaccine development [27]. Moreover, as seen with the recent GameStop trading event, Reddit is increasingly important in online conversations [28]. We note that Reddit and similar online sources are not necessarily representative of what the overall US general public feels [29]. However, Reddit provides insights on highly shared news, and can rapidly transmit both misinformation and accurate information [30, 31, 32].

It is important to understand the behavior of top users, how vaccine perceptions are related to events in the vaccine timeline and how vaccine discussion on Reddit migrates across subreddits that differ in their vaccine perceptions, as large-scale vaccination among the US general public expands. Most users of online platforms are passive or participate with a very low frequency. A small number of Reddit users are hyperactive and may over-proportionally influence vaccine perceptions online [33]. Thus, describing the behavior of hyperactive users is key to understanding shifts in vaccine perceptions. Understanding how perceptions are related to vaccine-related events may allow stakeholders to better design communication and education campaigns [34, 35] in response to vaccine distribution setbacks. Given the range of vaccine-related viewpoints online, greater insight on how discussions move across Reddit communities will allow stakeholders to better disseminate evidence-based information on Reddit. The purpose of this analysis was to detail the behavior of top Reddit users, posts’ relationship with events in the vaccine timeline, and the relationship between subreddits that shared COVID-19 vaccine posts.

## Methods

### Ethics statement

Our work did not require institutional review board approval as we used publicly accessible and deidentified posts from Reddit without any interaction with the posts’ authors. We did not use personally identifiable information, and paraphrased posts to reduce traceability.

### Data acquisition and processing

Using the Pushshift API and the Python Reddit API Wrapper [36, 37], we collected all posts on the entire Reddit, across all subreddits from January 1 2020 - December 14 2020 that contained both COVID-19 and vaccine keywords (see Supplement, only posts that had COVID-19 AND vaccine-related keywords were collected) derived from systematic reviews on the topic. We also collected metadata for each post e.g. the username, ID, subreddit. We then preprocessed our data as follows: 1) removed duplicate entries; 2) filtered out entries <50 characters as these generally do not provide enough information for meaningful analysis [38, 39]; 3) filtered the content using a curated set of search terms (as shown in the Supplement) to retain only COVID-19 vaccine-related content; 4) removed text in non-English languages, URLs, emojis, and punctuation.

### Hyperactive users

To better understand the possibly outsize influence of some individuals, we provided a descriptive overview of the behavior of top 10 users, focusing on content and number of posts.

### Topic modeling

We used topic modeling to understand changes in word prevalence within topics around COVID-19 vaccines (see Supplement for additional detail). Topic modeling is a computer-aided content analysis technique through which texts are organized into themes known as “topics” [40, 41]. We used an approach to topic modeling known as Structural Topic modeling (STM) [42, 43]. STMs [42, 43] enable the generation of topics with regards to document metadata such as date and source and other covariates relevant to the research question, such as new COVID-19 cases. We used the following metadata covariates for the STM model: date (1 was denoted for the first day and numbered sequentially after), new COVID-19 cases per day worldwide, new COVID-19 deaths per day worldwide (publicly available and both obtained from COVID-19 Data Repository by the Center for Systems Science and Engineering at Johns Hopkins University [44]), S and P 500 opening score (publicly available from the Wall Street Journal), post type (comment or post), score (upvotes - downvotes). We used worldwide cases and deaths instead of US cases/deaths as Reddit COVID-19 discussion centers on pandemic progression both globally and in the US, despite most users being from the US. These control variables may address underlying factors possibly influencing vaccine perceptions. By considering a broader picture of what may influence topic proportions around vaccine discussion, we can better test the claims relation to the association between specific events and topic proportions. March 11 2020 was denoted as the start date for our analysis, the date the World Health Organization declared COVID-19 a pandemic [45].

As STM is an unsupervised approach, the number of topics (k) to estimate is key to the analysis. We first estimated several models ranging from 5 to 30 topics. These models were then evaluated qualitatively by two authors (IS, AG) independently for 1) their ability to produce coherent topics and 2) appropriately capture topics regarding COVID-19 vaccination [46]. The two authors agreed on the same topic solution (k=20). Topic interpretation was influenced by authors’ first reading the top 100 most-cited COVID-19 peer-reviewed research articles and the top 10 most cited peer-reviewed research articles around topic modeling. Two authors assigned topics [Cohen’s kappa (k) >0.8] and a third author resolved disagreements when they arose.

We also detailed how events in the vaccine timeline (described in following section) were associated with topic prevalence. We generated linear regression models with expected topic proportions for each topic as dependent variables and vaccine events as main explanatory variables, with the following additional covariates: new COVID-19 cases per day worldwide, new COVID-19 deaths per day worldwide, SP 500 opening score, post type. To validate regression analyses in Figure 1, we undertook a close reading of the 100 most representative text fragments for exemplar topics. We found that these topics varied in line with the indicated events.

**Figure 1:**
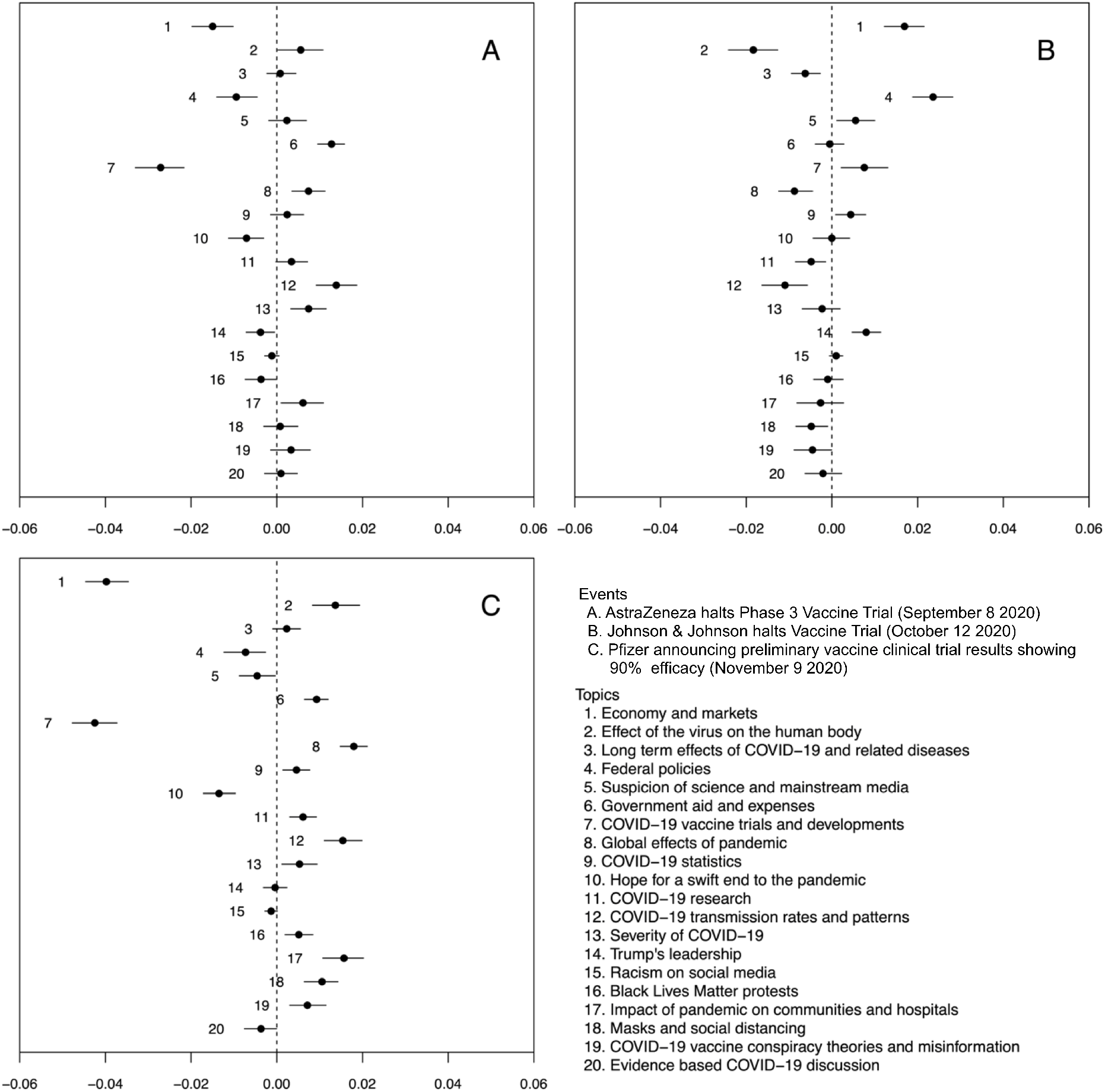
Values were generated from a regression where the outcome variable was the proportion of each document dedicated to each topic, given the selected STM model, and with various vaccine events as main explanatory variables. Topics on the right of the zero line were more likely to be brought up after the indicated event. Confidence intervals (95%) included both regression uncertainty and measurement uncertainty from the STM model.

### Selecting events of interest

We first assembled a preliminary list of COVID-19 vaccine-related events based on a review of online news sites and peer-reviewed vaccine research articles and consultation with experts on vaccination, resulting in a list of six events (see Supplement). We then conducted preliminary analyses with remaining events to determine the ones that were associated with the greatest shift in topic proportions for each topic. The three events below were selected as our final list, as these were associated with a shift in topic proportions for most topics. Events as follows: 1) AstraZeneca halts Phase 3 vaccine trial (September 8 2020); 2) Johnson Johnson temporarily halts vaccine trial (October 12 2020); 3) Pfizer announces preliminary vaccine clinical trial results showing 90% efficacy (November 9 2020).

### Social network analysis

Next, we conducted social network analysis to provide insights on how vaccine discussion on Reddit migrates across subreddits that differ in their vaccine perceptions, and the relationship between these subreddits [47]. While standard social networks tend to assess relationships between people, we used a network to describe relationships between subreddits, studying the connections between people as mediated by the subreddits they were in and the posts shared between these subreddits. We used node sizes to represent number of users in a subreddit, node edges to indicate shared COVID-19 vaccine posts between subreddits (an edge was indicated if there was >one shared post), and node labels to detail the subreddit name. Edge direction was based on whether a node had >50% of its posts made prior to its adjacent connecting node e.g. A->B if >50% of A’s shared posts were made before B. When analyzing the trajectory of posts from one subreddit to another, we assumed that posts moved from A->B->C if a post was made first in A, then followed by B and C. This may allow us to see how posts moved from one subreddit to another. We used the fast-greedy algorithm for cluster identification. We focused on the main social network in our data (largest component subgraph) and excluded all edges with a weight of one (i.e. all connections between subreddits that had only one post in common) and all clusters that had <15 vertices and whose vertices had a betweenness centrality <20 (we used a range of network characteristics to yield an easy to understand social network and the above measures yielded the clearest output).

## Results

Post-processing, we had 266,840 documents (25,400,556 words).

### Overview of hyperactive users

We reviewed the posts for the top 10 users who posted the most in our dataset, ranging from 159 - 278 posts/person. Six of these users posted evidence-based information (e.g. Effectiveness of the COVID-19 vaccine: real-world evidence from healthcare workers, Vaccine linked to reduction in risk of COVID-19 admissions to hospitals), but four users (one of these users was suspended from Reddit at time of writing) seemed to be skeptical of vaccination (e.g. Hell Gates says Vaccines are Americans’ only hope to return to Normal Life!, Doctors Around the World Issue Dire Warning: DO NOT get the experimental covid vaccine, At What Point Do We Realize Bill Gates Is Dangerously Insane?). Individuals skeptical of vaccination were common among those who posted the most frequently in our data.

### Topic modeling

Table 1 indicated the topics in the dataset, their proportions, and the top 10 words for each topic (see Table 1). Broadly, our data centered on the severity of the pandemic (Topic 13), hope for a swift end to the pandemic (Topic 10), and suspicion of science and mainstream media (Topic 5). The severity of the pandemic topic focused on death, risk and sickness in relation to the pandemic. The hope for a swift end to the pandemic topic was about hope, safety and the length of the pandemic. Finally, the suspicion of science topic was around reduced trust and belief in the media and science. We also noted several other topics, such as evidence-based COVID-19 discussion (exploring factually sound and true evidence about COVID-19) (Topic 20), COVID-19 transmission rates and patterns (Topic 12), the effect of the virus on humans (Topic 2), COVID-19 vaccine conspiracy theories and misinformation (e.g. Bill Gates-related vaccine conspiracy theories) (Topic 19), and racism on social media (Topic 15).

**Table 1:** Structural topic model results from 266,840 documents, March 11 2020 - December 14 2020, including the topic proportion and the top 10 words associated with each topic.

| Expected Topic Proportions | Topics Title | Top 10 words |
| --- | --- | --- |
| 0.1036 | Severity of COVID-19 | die, risk, life, normal, live, sick, yes, stop, stay, serious |
| 0.0941 | Hope for a swift end to the pandemic | long, term, hope, safe, next, available, wait, shot, pretty, rush |
| 0.0861 | Suspicion of science and mainstream media | believe, anti, tell, real, science, liter, bad, media, cure, trust |
| 0.0796 | Evidence based COVID-19 discussion | read, fact, understand, inform, clear, true, person, reason, post, evidence |
| 0.0702 | COVID-19 transmission rates and patterns | spread, population, herd, high, risk, rate, number, reduce, hospitalization, mortality |
| 0.0633 | Effect of the virus on the human body | common, system, response, human, influenza, body, cold, mutate, strain, similar |
| 0.0577 | Impact of pandemic on communities and hospitals | home, school, family, care, person, stay, live, learn, hospital, help |
| 0.0523 | Global effects of pandemic | world, country, public, global, travel, economy, social, govern, open, state |
| 0.0500 | COVID-19 vaccine trials and developments | data, phase, trial, clinic, safety, studies, drug, efficacy, severe, receive |
| 0.0478 | COVID-19 statistics | death, million, rate, dead, season, die, total, number, second, near |
| 0.0381 | Economy and markets | market, companies, stock, product, industry, supply, price, sell, demand, billion |
| 0.0376 | Federal policies | trump, president, nation, state, elect, administration, federal, house, unit, response |
| 0.0359 | Government aid and expenses | govern, money, free, pay, business, cost, spend, economy, system, support |
| 0.0325 | Long term effects of COVID-19 and related diseases | medical, damage, cancer, doctor, heart, polio, measles, harm, medicine, child, blood |
| 0.0310 | Black Lives Matter protests | game, watch, video, police, fire, sport, street, post, show, red |
| 0.0309 | COVID-19 vaccine conspiracy theories and misinformation | bill, world, power, human, war, russia, mark, control, chip, conspiracy |
| 0.0294 | COVID-19 research | research, link, studies, found, scientific, respiratory, science, paper, associate, article |
| 0.0291 | Masks and social distancing | wear, mask, social, protect, spread, face, public, hand, person, distance |
| 0.0160 | Racism on social media | chance, please, remember, pass, remove, black, message, thank, stick, attend |
| 0.0148 | Trump's leadership | march, trump, perfect, control, disappear, march, anybody, fine, false, great |

Note: The topic proportions indicated the proportion of the corpus that belongs to each topic.

We then explored how various events in the vaccine timeline were related to topic prevalence (see Figure 1). We observed an association between AstraZeneca temporarily halting its vaccine trials, and increased discussion around government expenses, such as funds spent on businesses, and supporting the economy. Similarly, we found an association between Johnson and Johnson temporarily halting its vaccine trial, and increased discussion of federal policies and then-US President Donald Trump’s leadership. We found an association among Johnson and Johnson temporarily halting its vaccine trial and greater discussion around suspicion of science and mainstream media. Similarly, there was an association between Pfizer announcing preliminary Phase 3 results showing 90% vaccine efficacy and reduced discussion about suspicion of science and mainstream media. We detailed an association between Johnson and Johnson temporarily halting its vaccine trial and reduced discussion around COVID-19 vaccine conspiracy theories and misinformation. We found an association between Pfizer announcing preliminary vaccine clinical trial results, an increase in discussion around COVID-19 vaccine conspiracy theories and misinformation, and a corresponding decrease in evidence-based COVID-19 discussion. We also found that new COVID-19 deaths and cases were positively associated with increased discussion around COVID-19 vaccine conspiracy theories and misinformation, and suspicion of science and mainstream media, highlighting the relationship between COVID-19 progression and similar rises in misinformation (See Supplement for full results).

### Subreddit networks

To understand the relationship between subreddits that shared COVID-19 vaccine posts, we analyzed the greatest component subgraph, as this was substantially larger than all other subgraphs which had 1-5 nodes and did not provide for meaningful conclusions (see Figure 2). The largest node/subreddit (r/Coronavirus, the official community for COVID-19 on Reddit) had 2.4 million users. Nodes connected by an edge shared two to 41 posts.

**Figure 2:**
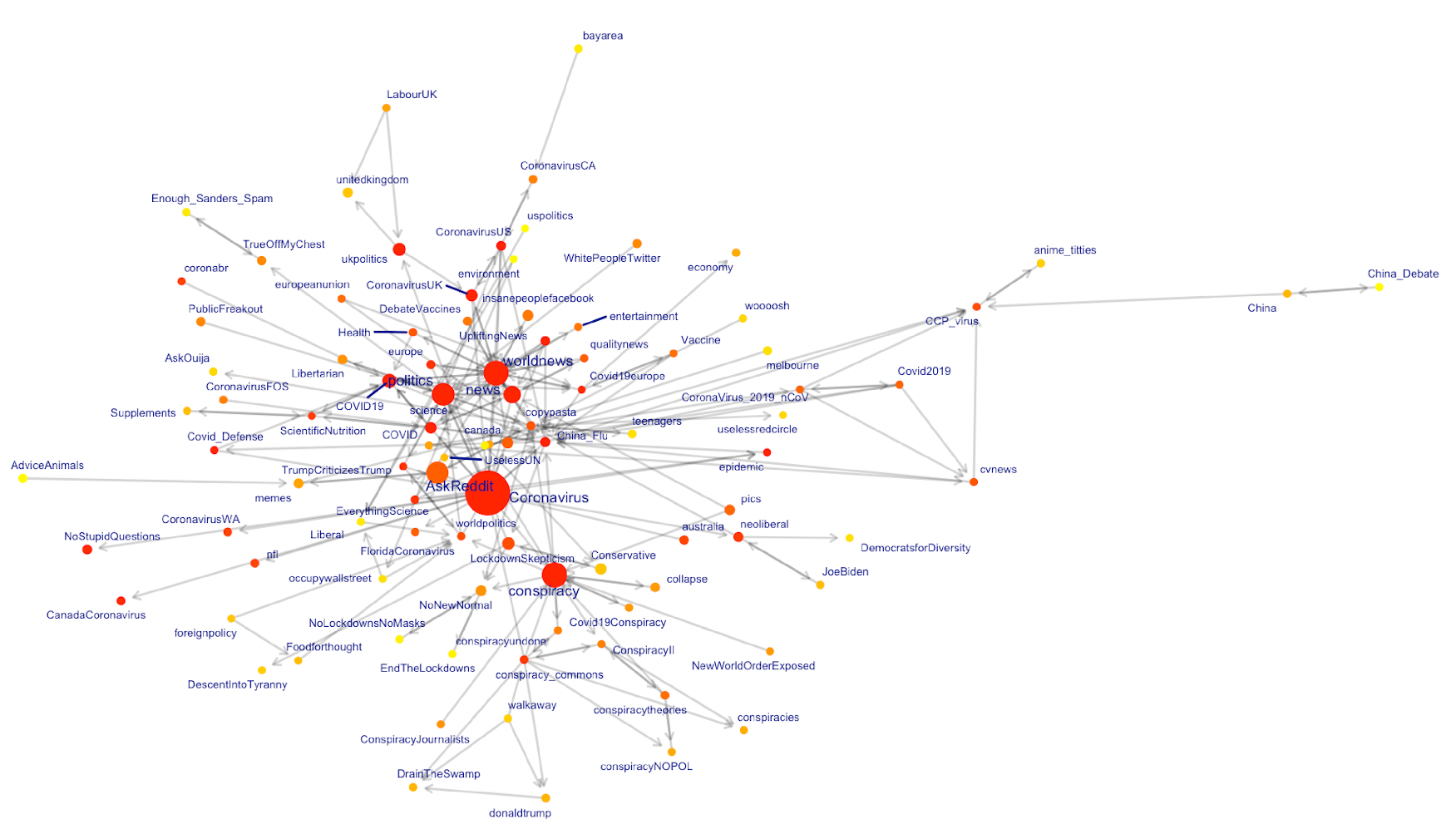
The largest subgraph above shows subreddits that have COVID-19 vaccine-related posts. Node sizes represent subreddit user sizes, node edges indicate shared COVID-19 vaccine posts between subreddits, and node labels indicate the subreddit name. Node color is based on centrality (red is higher centrality, yellow is lower centrality). Edge direction was based on whether a node had >50% of its posts prior to its adjacent connecting node e.g. A->B if >50% of A’s shared posts were made before B. We used larger labels for subreddit nodes with >5,000 COVID-19 vaccine posts.

We found nine posts that were first posted in r/Coronavirus and then subsequently posted in at least one subreddit. Posts were reposted one to 10 times. Eight of these posts concerned evidence-based information (e.g. COVID-19 timeline, Vaccine development timeline) and were reposted in other subreddits favoring evidence-based information (e.g. AmericanPolitics, worldnews). However, one post (COVID-19 much milder than believed) was aligned with vaccination skepticism and subsequently posted in subreddits favoring vaccine skeptic narratives (e.g. conspiracy, LockdownSkepticism - we read through the first 50 posts in these subreddits and verified they were largely around disagreement with evidence-based measures to mitigate the pandemic). This suggests that misinformation is present in some subreddits which generally feature accurate information. This may also indicate that most posts which start in the main COVID-19 subreddit (r/Coronavirus) and then re-posted in other subreddits tend to be evidence-based. However, a minority of posts in r/Coronavirus are skeptical of vaccination, but then do not get reposted in evidence-based subreddits, but instead in subreddits broadly skeptical of vaccination.

## Discussion

Our analysis of 266,840 posts on COVID-19 vaccines between March 11 2020 - December 14 2020 generated several key findings. First, there was a relationship between interim positive announcements followed by increased vaccine misinformation, and a relationship between halting vaccine trials and reduced misinformation discussion. Past research has indicated shifts in vaccine perceptions with time [18, 19, 48]. We expand on that work, suggesting an association between events in the vaccine timeline and vaccine perceptions. Information skeptical of vaccination may flow from a regulated and legitimate source to avenues centering on misinformation and distrust in science. Previous research indicated how antivaccine posts travel online, with users largely moving from one antivaccine post to another [49, 50]. Building on this work, we propose that individuals skeptical of vaccination may selectively highlight posts from legitimate online environments and then forward these posts in arenas aligned with vaccine-skeptic narratives, moving information that was previously under the purview of a more neutral, science-trusting audience to individuals skeptical of vaccination - perhaps providing opportunities to engage such individuals and reduce misinformation. The strength of our work is the use of computational methods to explore how Reddit vaccine perceptions are associated with events in the vaccine timeline and how posts move among environments with differing vaccine perceptions. Such outcome measurement is central to understanding how vaccine perceptions shift, allowing for accurate public health messaging capable of improving COVID-19 vaccine perceptions.

There was an association between positive vaccine developments and an increase in discussion of COVID-19 vaccine misinformation, and a relationship between development setbacks and reduced misinformation discussion. Past research has indicated shifting vaccine perceptions over time [5], but there is limited research on specific events, especially vaccine trials and their relationship with vaccine perceptions. Previous work also indicated that COVID-19 misinformation can be remedied with scientific facts [6], but we highlight the complexity of the phenomenon. The spread and production of misinformation can sometimes be due to confirmation bias, where individuals consume, interpret, and favor information that supports their beliefs [51]. For true antivaxxers, COVID-19 vaccine successes may be interpreted as attempts by Bill Gates to track the population through microchips, and thus news around vaccine successes may be interpreted in a misinformation framework, perhaps explaining the relationship between vaccine success and increased discussion around COVID-19 vaccine conspiracy theories. Similarly, when vaccine trials are halted, such news may cohere with antivaxxers, who may have no interest in engaging with news that possibly demonstrates the failure of medical science - given antivaxxers’ distrust of medical experts [52], perhaps explaining the reduced misinformation discussion. Thus, simply presenting scientific data to antivaxxers [6] may not be effective, as demonstrated in a study where presenting some antivaxxers with facts made them more antivaccine [53].

There was a relationship between a vaccine trial halting and increased discussion around suspicion of science and mainstream media, and a vaccine trial being effective and reduced discussion around suspicion of science and mainstream media. Factors such as political conservatism and lower levels of education may be associated with lack of trust in science [54], and we build on such research by suggesting that news around science successes and setbacks is associated with trust in science. In an environment where individuals are unsure what to believe around vaccines [55], we propose that vaccine successes build faith in science, and vaccine setbacks erodes this trust.

We also documented how posts skeptical of vaccination may move from more legitimate avenues to arenas where vaccine-skeptic narratives are more popular. In addition, such posts were popular among some highly active users in our dataset. COVID-19 misinformation is present in mainstream environments and does not always get fact-checked [56] and Reddit is no different. Individuals with largely antivaccine beliefs seek out information that coheres with their views [49]. We build on this work and suggest that individuals skeptical of vaccination also look for information from venues that tend to have evidence-based discussion, but then may interpret such information in line with their views and moral foundations, later sharing such information in forums more skeptical of vaccination. This may indicate that skeptics of vaccination do venture out of their echo chambers to enter spaces where accurate information is the norm - presenting attractive opportunities for constructive intervention.

To improve COVID-19 vaccine perceptions, minimize misinformation, and increase vaccination rates, public health authorities should conduct tailored interventions and communications campaigns to counter the rhetoric of vaccine misinformation [57, 58]. An example intervention could ask respondents to determine information accuracy around vaccines [59, 60] nudging individuals through the design of these programs toward accurate vaccine information. It is possible that interventions of this sort could shift the beliefs of the vaccine hesitant and thereby boost vaccine uptake, despite potentially little or no effect on committed opponents of vaccination. The concomitant spread of misinformation about COVID-19 vaccines and scientific implications provides insights about the mechanism of misinformation spread. Given our findings around a vaccine trial halting and increased discussion around suspicion of science, we suggest that scientists be more communicative on the difficulties they face in creating vaccines to mitigate science mistrust. Communications campaigns can harness these findings and forward evidence-based posts in subreddits where misinformation is common, when vaccine trial data is released. Given the possibility that individuals seemingly more interested in antivaccine narratives may sometimes venture into more evidence-based environments, interventions can target skeptics or critics of vaccination who sometimes enter more mainstream spaces, engaging them with more evidence-based information, keeping in mind how antivaxxers may deal with such information. Similarly, as legitimate online spaces contain COVID-19 vaccine misinformation, more effective moderation policies can be enacted in these and similar environments e.g. perhaps including a “verified” tag to a post if it comes from a credible source. Such measures may augment health outcomes through several modes. For example, improved vaccine perceptions may lead to reduced vaccine hesitancy and thereby increase vaccine acceptance and COVID-19 vaccination rates. Reduced vaccine misinformation may also improve trust in science and health systems, more broadly, enhancing larger efforts to address health disparities observed in vaccination coverage and many other areas [61].

## Limitations

Our findings relied on the validity of data collected with our search terms. We searched all of Reddit for COVID-19 vaccine posts, and our data contained text fragments representative of vaccine perceptions. We are thus confident in the comprehensiveness of our data. Any use of Reddit data presents several challenges and limitations. As no personal information is collected on Reddit, the demographic makeup of users is unknown [62]. Some evidence suggests that Reddit users are likely to be male, younger than the general population and mostly based in the US [63]. Our results should be interpreted in line with this probable gender and age skew of our data.

It was not possible to determine what posts were viewed by skeptics of vaccination in more legitimate subreddits, but subsequently not reposted in subreddits more supportive of anti-vaccine narratives, thereby providing more support for our suggestion around confirmation bias. It is possible that posts were made in one subreddit before another purely due to chance, and that the directionality assumed is due to coincidence. We cannot be certain why individuals created the text in our data, the processes behind the shift in narratives, and why individuals shared the same post in more than one subreddit, and we cannot address these mechanisms with our data. Future work can address these questions and explore the motivations of those creating and sharing such text. We conducted a retrospective and observational study, and thus cannot draw causal conclusions regarding vaccine perceptions. It is possible that other vaccine-related events may have caused the observed changes, and that vaccine success stimulated debate that brought to the surface existing antivaccine discussion, instead of causing it.

## Conclusion

There was a relationship between vaccine efficacy and increased discussion around vaccine misinformation, and an association between halting of vaccine trials and reduced misinformation discussion. Posts skeptical of vaccination can move from more legitimate avenues to arenas where antivaccine narratives are more popular, suggesting that individuals skeptical or opposed to vaccines do venture out of their online echo chambers, providing potential opportunities to engage such individuals and reduce misinformation. Given the time period of our data extends to the period immediately prior to the launch of large-scale vaccination among the US public, stakeholders may utilize our findings to better design and inform upcoming vaccination communication efforts.

## Data Availability

Data is available from authors' at reasonable request.

## Availability of data and materials

Data is available from authors’ at reasonable request.

## Competing interests

The authors declare that they have no competing interests.

## CrediT authorship contribution statement

All authors made significant contributions to the manuscript. The following were the respective roles for each author: NK, AG, CM, IC, KJ, MH, NY, NH, SNS, KK, SO contributed to the study design, hypothesis generation, data collection, data analysis, data interpretation, and manuscript write-up and review. MDC, JDT, OP, WT, CB, AT, JLS, SMJ, SAM contributed to the manuscript write-up and review.

## Acknowledgements

Study was funded by the Yale Institute for Global Health and the Whitney and Betty MacMillan Center for International and Area Studies at Yale University. The funding bodies had no role in the design, analysis or interpretation of the data in the study. This study was pre-registered on the Open Science Framework (osf.io/urp2a).

## Supplement

### Software

All analysis was conducted using python and R with the following packages: datetime [64], dplyr [65], ggraph [66], grid [67], gridExtra [68], igraph [69], lubridate [70], NumPy [71], pandas [72], pracma [73], praw [74], quanteda [75], readtext [76], readr [77], stm [43], stminsights [78], splines [79], stringr [80], textclean [81], tidygraph [82], tidytext [83], tidyverse [84].

### Search terms

#### COVID-19 keywords

(coronavirus OR coronaviruses OR corona virus OR corona viruses) OR (coronavirus infections OR corona virus infections) OR ’(betacoronavirus OR beta coronavirus OR beta coronaviruses OR betacoronaviruses OR beta corona virus OR beta corona viruses OR betacorona virus OR betacorona viruses) OR (severe acute respiratory syndrome coronavirus OR severe acute respiratory syndrome corona virus) OR SARS CoV-2 OR cov2 OR sars 2 OR COVID OR (coronavirus 2 OR corona virus 2) OR covid19 OR nCov OR (new coronavirus OR new corona virus) OR (novel coronavirus OR novel corona virus) OR (novel coronavirus pneumonia OR novel corona virus pneumonia) OR ncp OR (pneumonia AND (wuhan—china—chinese—hubei))

#### Vaccine keywords

(vaccine OR vaccinate OR vaccinated OR vaccinating OR vaccines OR vaccinates OR vaccination OR vaccinations) OR (immunisation OR immunise OR immunising OR immunisations OR immunises OR immunised) Or (immunization OR immunizations OR immunize OR immunized OR immunizes OR immunizing)

### Online news sites

historyofvaccines.org/content/articles/coronavirustimeline

mmunize.org/timeline

biospace.com/article/a-timeline-of-covid-19-vaccine-development

fortune.com/2020/12/30/covid-vaccine-first-coronavirus-cases-timeline-2020

### Vaccination experts

We identified key scholars in vaccination through the number of articles (>10) published regarding vaccination. We then contacted the identified researchers and asked them to assist.

### Longlist of vaccine-related events

Fauci says he is cautiously optimistic that a vaccine will be effective and achieved within 1 or 2 years (May 12 2020)

United States and AstraZeneca Form Vaccine Deal (May 21 2020)

Moderna Vaccine Begins Phase 3 Trial, Receives $472M From then-US President Donald Trump’s Administration (July 27 2020)

AstraZeneca Halts Phase 3 Vaccine Trial (September 8 2020)

Johnson Johnson Halts Vaccine Trial (October 12 2020)

Pfizer announcing preliminary vaccine clinical trial results showing 90% efficacy (November 9 2020)

### Topic modeling

Within topic modeling, a topic is a distribution over a vocabulary [85]. For example, in a topic denoted “vape”, there is likely a greater probability that the terms “smoke” and “device” occur than the words “peanut” and “tomato”. “Smoke” may appear in both “vape” and “cooking” topics with different contextual meanings. Given the topic is a distribution, “smoke” may appear with other high-probability terms like “roast” and “fry” in the “cooking” topic, but with terms like “nicotine” and “device” in the “vape” topic. Thus, topics can be understood as if a person was to talk about a topic and when doing so, tended to use some words than others when the topic is “cooking” compared to “vape”. Topic models are apt for analyzing large quantities of textual data via an automated technique for providing context.

The key innovation of STM is that it can incorporate metadata or information about each document. This allows metadata covariates, such as new COVID-19 cases per day, to influence topic discovery. Metadata can affect both topic prevalence and content. Metadata covariates for topical prevalence allow the metadata to affect topic frequency. Similarly, covariates in topical content allow the metadata to affect the word rate within a topic or how a topic is discussed [43]. The STM process will output documents and vocabulary for analysis [43]. Output can be investigated in a range of ways, such as detailing words associated with topics or the relationship between metadata and topics. Model output can be used to conduct hypothesis testing around these relationships.

The number of topics was based on our understanding of the dataset and how other researchers interpreted STM results [46, 86]. Choosing the number of topics was also influenced by post-estimation validation outcomes and past work [46]. As per standard content analysis [87], topic model validation also needs qualitative review, where researchers assess the interpretability and relative efficacy of models based on their subject matter expertise and data context. Our final model [k=20] provided the greatest external validity and most semantically coherent output of distinctive topics. Above the indicated number of topics, there were diminishing returns for solutions, as the substantive meaning and coherence of categories started to break down. Below the indicated number of topics, variation decreased and specific topics got placed into more generic categories. Validating a topic model is not the same as evaluating a statistical model regarding a population sample [88]. The goal is to identify the framework which best describes the data, not estimating population parameters [88].

Most of the text was produced and consumed by people who were interested in the COVID-19 vaccine, and this lens was used to interpret the presence/absence of topics and words. Most of the topic labels were straightforward and did not require much interpretation. To characterize topics in the COVID-19 vaccine narrative, we qualitatively coded each topic by investigating word clouds based on each topic and reviewing exemplar documents which detailed high proportions of each topic [85]. The topic we classified as “Economy and markets” had the following most frequently occurring words: market, company, stock, product, industry, supply, price, sell, demand, billion, economy, trade, million, invest, high. Exemplar documents which exhibited high proportions of this topic indicated a preoccupation with these words. Thus, the interpretation of the topic was clear, given the genre of the narrative and relying on research regarding prominent topics around the COVID-19 vaccine.

Topic validation is key to assessing whether the substantive meaning of the topic and its words are parallel with the qualitative meaning of the text and we used methodological guidance from past research for this purpose [85, 41]. Past work advocated the use of sample documents to validate each topic’s substantive meaning. Determining the number of sample documents to use is based on the amount of resolution needed by a social scientist to answer the research question using topic modeling methods [89]. Thus, determining the number of sample documents is a largely qualitative process, dependent on the research question at hand. To determine the appropriate number of documents to sample, we searched the social science literature for studies that used topic modeling, based on the following study characteristics: 1) similar research questions as our study; 2) similar topic areas as our study; 3) study data was drawn from similar sources as our study. We searched databases such as Web of Science Core Collection, Embase, PsycINFO, MEDLINE and Sociological Abstracts. We used keywords such as vaccine, misinformation, and topic modeling. The paper by Farrell (2016) was determined to be most similar to our study based on the assessed characteristics. Farrell (2016) explored ideological polarization in climate change and used a broad range of sources, such as press releases, published papers, and website articles. Based on the nature of the research question and large range of sources, Farrell (2016) determined that a sample of 50 documents was sufficient to validate the substantive meaning of the topic output. Given the similarities between Farrell’s (2016) study and ours on a range of characteristics, we similarly determined that a sample of 50 documents was adequate to validate the topics. We used findThoughts and plotQuote within the STM package to examine the top 50 associated documents for each topic to validate a topic’s substantive meaning. Determination of the top 50 documents was based on ranking topics by the maximum a posteriori estimate of the topic’s theta value, which represents the modal estimate of the proportion of word tokens assigned to the topic with the model. These top 50 documents were read by two of the authors to determine validity (k>0.8). A third author resolved disagreements where necessary.

### Regression results for models underlying Figure 1

**Supplementary Table 1:** Regression output for difference in topic proportions before and after AstraZeneca halted Phase 3 vaccine trials (September 8 2020)

| Topic | (1) | (2) | (3) | (4) | (5) | (6) | (7) | (8) | (9) | (10) | (11) | (12) | (13) | (14) | (15) | (16) | (17) | (18) | (19) | (20) |
| --- | --- | --- | --- | --- | --- | --- | --- | --- | --- | --- | --- | --- | --- | --- | --- | --- | --- | --- | --- | --- |
| (Intercept) | -3.294e-02** | 1.293e-01*** | 5.171e-02*** | 2.670e-02* | 8.675e-02*** | 5.019e-02*** | -6.125e-02*** | 4.684e-02*** | 9.726e-02*** | 1.07e-2 | 7.712e-02*** | 1.171e-01*** | 1.172e-01*** | 5.679e-02*** | 2.068e-02*** | 5.77e-3 | 4.821e-02*** | 8.585e-02*** | 1.96e-2 | 4.618e-02*** |
| Score | 6.38e-7 | 4.21e-6 | 2.75e-7 | 9.910e-06*** | -4.92e-7 | -1.50e-6 | 8.548e-06*** | -1.39e-6 | -4.143e-06** | -2.15e-6 | 2.15e-6 | -8.309e-06*** | -1.464e-05*** | 9.591e-06*** | -1.331e-06* | 6.464e-06*** | 1.90e-6 | -3.833e-06** | 1.23e-6 | -7.153e-06*** |
| New deaths | -1.60e-7 | -1.237e-06*** | -8.41e-8 | 7.613e-07*** | 3.29e-7 | 4.833e-07** | -2.978e-06*** | 5.555e-07** | -2.09e-8 | -8.072e-07*** | 3.04e-7 | -2.05e-7 | -1.36e-7 | 1.39e-7 | 5.58e-8 | 9.847e-07*** | 5.760e-07* | 9.788e-07*** | 6.691e-07** | -2.02e-7 |
| New cases | 5.32e-9 | -8.652e-09** | 1.07e-9 | -7.177e-09** | 6.318e-09* | -5.99e-10 | 3.62e-10 | -1.87e-9 | 2.18e-9 | -6.216e-09* | -4.925e-09** | 6.103e-09* | 3.45e-9 | 1.48e-9 | -6.23e-11 | -1.86e-9 | 2.88e-9 | 1.29e-9 | 3.71e-9 | -3.01e-9 |
| Post type | 2.329e-02*** | -2.749e-02*** | -7.265e-03*** | 2.159e-02*** | -1.142e-02*** | -2.11e-3 | 1.392e-02*** | 7.578e-03** | -2.095e-02*** | -4.256e-02*** | 2.30e-3 | -3.433e-02*** | -4.688e-02*** | 4.193e-02*** | 1.017e-02*** | 2.335e-02*** | 1.899e-02*** | -3.78e-3 | 3.497e-02*** | -1.22e-3 |
| S and P 500 opening score | 2.845e-05*** | -1.01e-5 | -8.161e-06** | 7.01e-6 | -1.106e-05* | -1.401e-05*** | 4.506e-05*** | -1.12e-6 | -4.33e-6 | 2.126e-05*** | -1.537e-05*** | -1.51e-6 | -1.392e-05*** | -6.52e-6 | -1.55e-6 | 8.337e-06** | -5.07e-6 | -1.977e-05*** | -3.18e-6 | 5.58e-6 |
| s(days)1 | 3.011e-02*** | -2.129e-02*** | 6.52e-4 | 1.373e-02** | -8.818e-03* | -2.06e-3 | 3.652e-02*** | 1.656e-02*** | -4.112e-02*** | 1.987e-02*** | 2.21e-3 | -5.089e-02*** | -2.351e-02*** | 1.418e-02*** | 6.725e-03*** | 6.681e-03* | 2.50e-3 | -2.450e-02*** | 2.863e-02*** | -6.29e-3 |
| s(days)2 | -1.484e-02*** | -5.21e-3 | 4.929e-03* | -1.799e-02*** | 1.180e-02*** | 1.035e-02*** | 2.092e-02*** | 7.352e-03** | -2.312e-02*** | 1.150e-02*** | 2.30e-3 | -2.302e-02*** | 2.896e-02*** | -1.847e-02*** | -6.703e-03*** | -1.351e-02*** | 5.81e-4 | -1.544e-02*** | 1.893e-02*** | 2.061e-02*** |
| s(days)3 | 1.414e-02*** | -4.835e-02*** | 6.067e-03* | 6.21e-3 | 1.344e-02*** | 1.566e-02*** | 5.31e-3 | 6.457e-03* | -3.753e-02*** | 5.21e-4 | 3.60e-3 | -5.933e-02*** | 6.504e-03* | -1.719e-02*** | 6.410e-03*** | 1.404e-02*** | 2.922e-02*** | 4.45e-3 | 2.320e-02*** | 7.064e-03** |
| s(days)4 | -6.97e-4 | -1.897e-02*** | 9.194e-03*** | -1.901e-02*** | 2.546e-02*** | 1.500e-02*** | 1.448e-02*** | -8.509e-03** | -3.913e-02*** | 1.830e-02*** | 1.13e-3 | -5.989e-02*** | 1.206e-02*** | -1.503e-02*** | 6.60e-4 | -2.12e-3 | 1.791e-02*** | 2.169e-02*** | 2.206e-02*** | 5.40e-3 |
| s(days)5 | 4.15e-3 | -4.147e-02*** | 1.211e-02*** | 1.108e-02* | 2.953e-02*** | 2.189e-02*** | 1.775e-02*** | -3.23e-3 | -4.646e-02*** | 1.194e-02** | 6.14e-3 | -7.955e-02*** | 2.09e-3 | -1.282e-02*** | 5.482e-03*** | 1.24e-3 | 1.751e-02*** | 3.83e-3 | 3.022e-02*** | 8.379e-03* |
| s(days)6 | -5.345e-02*** | -2.651e-02*** | 1.428e-02** | -8.07e-3 | 2.737e-02*** | 3.677e-02*** | -5.278e-02*** | 9.991e-03* | -2.298e-02*** | 1.40e-3 | -1.49e-3 | -2.554e-02*** | 3.529e-02*** | -1.050e-02* | -2.62e-3 | -1.69e-3 | 3.237e-02*** | 2.215e-02*** | 1.406e-02* | 1.205e-02* |
| s(days)7 | 1.11e-2 | -2.071e-02** | 8.33e-4 | 2.081e-02*** | 1.809e-02*** | 3.605e-02*** | 3.36e-3 | 1.38e-4 | -4.663e-02*** | 7.53e-3 | 8.11e-3 | -6.400e-02*** | 6.10e-3 | -1.527e-02*** | 2.53e-3 | -6.11e-3 | 8.93e-3 | 6.21e-3 | 1.999e-02*** | 3.01e-3 |
| s(days)8 | -2.862e-02*** | -1.560e-02* | 1.782e-02*** | -3.137e-02*** | 2.988e-02*** | 3.502e-02*** | -7.56e-3 | -5.38e-3 | -4.277e-02*** | 3.182e-02*** | 2.20e-4 | -5.346e-02*** | 2.792e-02*** | -2.000e-02*** | 2.21e-3 | -1.182e-02* | 3.946e-02*** | 1.112e-02* | 1.179e-02* | 9.30e-3 |
| s(days)9 | -3.770e-02*** | -1.17e-2 | 2.908e-02*** | -6.99e-3 | 2.225e-02*** | 3.134e-02*** | 1.04e-2 | -7.65e-3 | -4.539e-02*** | 1.929e-02*** | 1.023e-02* | -5.944e-02*** | 1.572e-02* | -2.044e-02*** | 5.742e-03** | -1.060e-02* | 1.832e-02** | 4.76e-3 | 1.432e-02* | 1.847e-02** |
| s(days)10 | -3.324e-02*** | -1.12e-2 | 2.200e-02*** | -1.08e-2 | 2.482e-02*** | 3.259e-02*** | -2.04e-3 | -1.02e-2 | -4.499e-02*** | 2.402e-02*** | 4.83e-3 | -5.915e-02*** | 2.026e-02** | -1.950e-02*** | 3.03e-3 | -1.034e-02* | 3.459e-02*** | 8.34e-3 | 1.339e-02* | 1.370e-02* |
| timePre | -1.503e-02*** | 5.678e-03* | 7.91e-4 | -9.514e-03*** | 2.36e-3 | 1.274e-02*** | -2.716e-02*** | 7.393e-03*** | 2.46e-3 | -7.032e-03*** | 3.35e-3 | 1.386e-02*** | 7.466e-03*** | -3.821e-03* | -1.15e-3 | -3.648e-03* | 6.158e-03* | 8.22e-4 | 3.31e-3 | 1.04e-3 |

**Supplementary Table 2:**
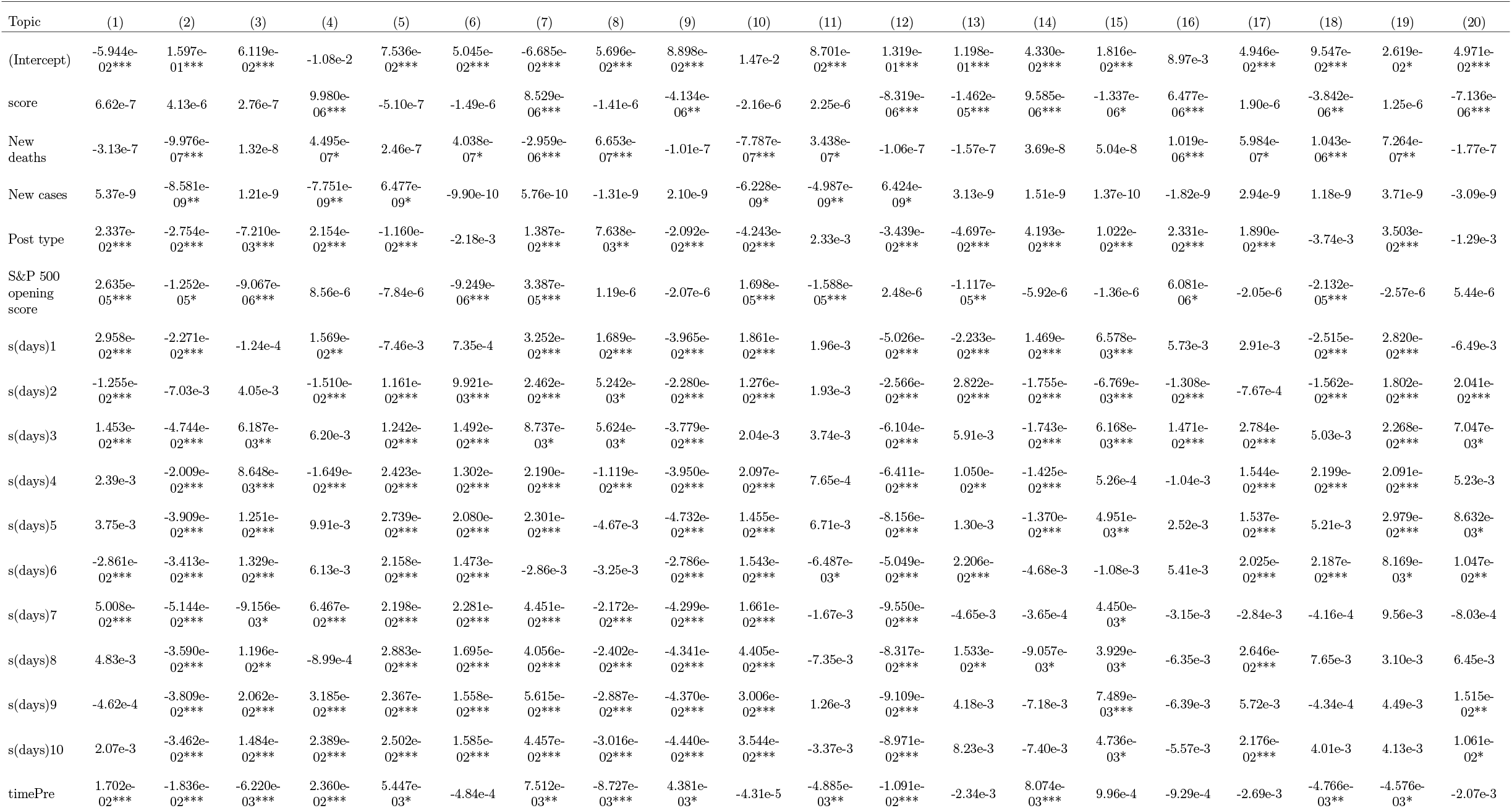
Regression output for difference in topic proportions before and after Johnson & Johnson halted vaccine trials (October 12 2020)

**Supplementary Table 3:**
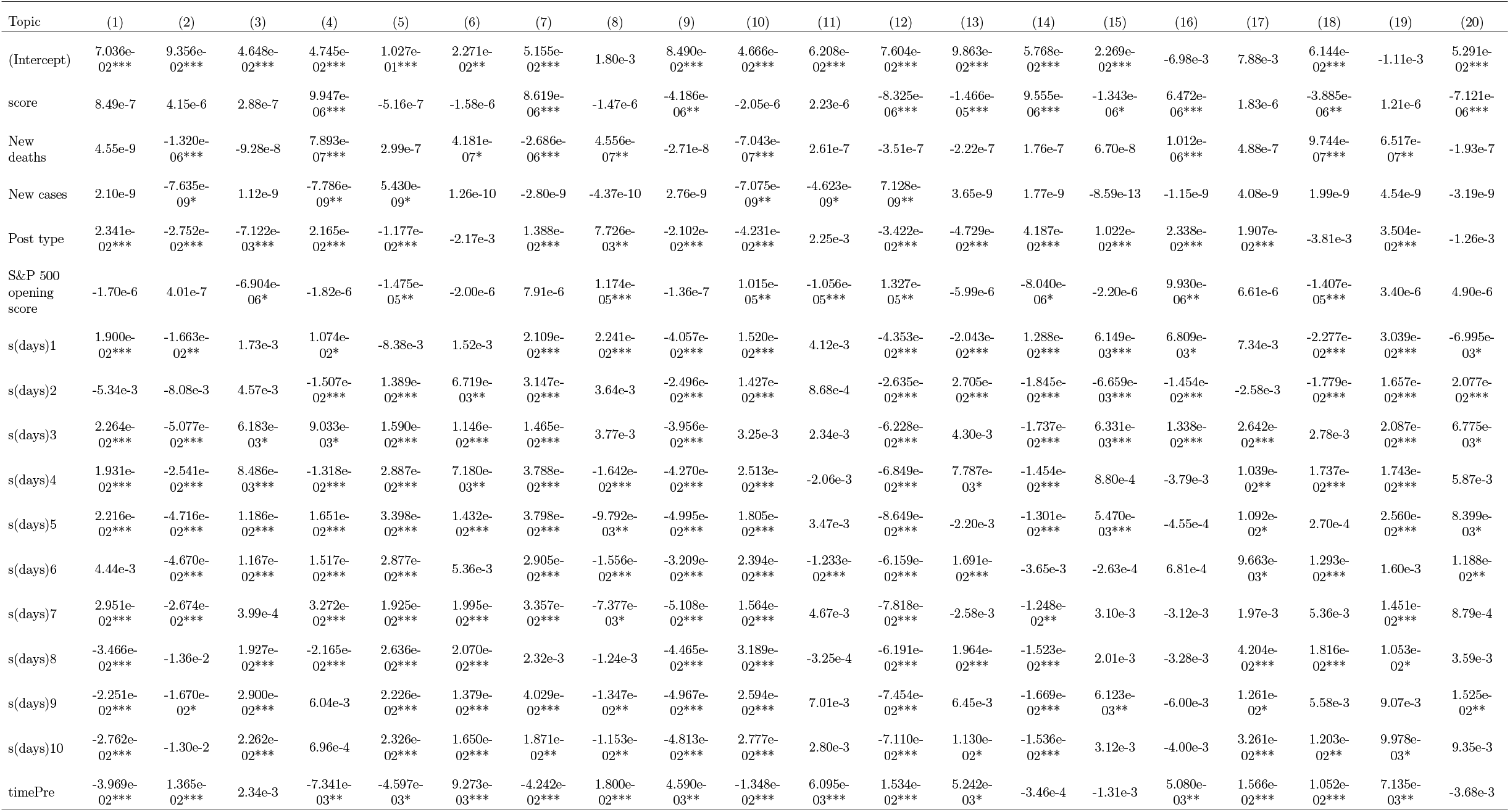
Regression output for difference in topic proportions before and after Pfizer announced 90% vaccine efficacy (November 9 2020)

## Highlights

Association between vaccine trials and discussion on vaccine misinformation.

Vaccine discussion moves from legitimate avenues to antivaccine arenas.

Findings have implications for development of vaccine perception interventions.

Interventions may nudge individuals toward accurate information.

## Notes

### Competing Interest Statement

The authors have declared no competing interest.

### Author Declarations

This study did not involve human subjects.

